# Cardiovascular autonomic dysfunction precedes cardiovascular disease and all-cause mortality: 11-year follow-up of the ADDITION-PRO study

**DOI:** 10.1101/2024.12.18.24319131

**Authors:** Jonas R. Schaarup, Lasse Bjerg, Christian S. Hansen, Erik L. Grove, Signe T. Andersen, Dorte Vistisen, Søren Brage, Annelli Sandbæk, Daniel R. Witte

**Affiliations:** Department of Public Health, Aarhus University, Aarhus, 8000, Denmark; Steno Diabetes Centre Aarhus, Aarhus, 8200, Denmark; Steno Diabetes Centre Copenhagen, Herlev, 2730, Denmark; Department of Clinical Medicine, Faculty of Health, Aarhus University, Aarhus, 8200, Denmark; Department of Cardiology, Aarhus University Hospital, 8200, Aarhus, Denmark; Department of Medicine, Gødstrup Hospital, Herning, Denmark; Novo Nordisk A/S, Søborg, Denmark; MRC Epidemiology Unit, University of Cambridge, United Kingdom

**Keywords:** Week-long heart rate variability, hourly heart rate variability, autonomic dysfunction, heart failure, all-cause mortality, high risk of diabetes, ADDITION-PRO study

## Abstract

**Background:** Cardiovascular autonomic dysfunction remains a silent complication in people at high risk of diabetes. We aim to determine the impact of week-long heart rate variability (HRV) on the risk of cardiovascular events and mortality in this population.

**Methods:** Week-long HRV and mean heart rate (mHR) were measured in 1,627 participants from the ADDITION-PRO study between 2009-2011. As measurement for HRV, we calculated a proxy for standard deviation of normal heartbeat (SDNN) both weekly, daily and hourly. Data on cardiovascular events (CVD) and all-cause mortality were obtained from Danish patient registers until 2021. We fitted poisson regression to determine incidence rate ratios (IRR) for major adverse cardiovascular events (MACE) (myocardial infarction, stroke, cardiovascular death), heart failure, and all-cause mortality.

**Results:** Mean (SD) age was 66 years (7), and 47 % were women. The population had a mean (SD) week-long SDNN of 139.0 (32.3) ms. Week-long HRV index SDNN showed an IRR of 0.82 (CI: 0.69; 0.97), 0.76 (CI: 0.58; 0.99), and 0.79 (CI: 0.66; 0.94) per SD for MACE, heart failure, and all-cause mortality, respectively. The risk for MACE, heart failure, and all-cause mortality was higher at SDNN values below 120ms. SDNN measurements taken from 6:00-7:00 AM showed the strongest association with the risk of MACE. Lower SDNN was consistently associated with higher all-cause mortality risk across all hours of the day. Adjustment for concurrent physical acceleration and heart rate did not materially change the magnitude of these associations. Night-time heart rate was associated with a higher risk of MACE, heart failure and all-cause mortality.

**Conclusion:** Cardiovascular autonomic dysfunction, measured by week-long HRV, is associated with higher risk of CVD, heart failure and all-cause mortality. Certain time frames of the day for HRV and heart rate under free-living conditions showed higher risk of CVD. Hence, long-term HRV and the diurnal response are linked with CVD risk among people with high risk of diabetes. Studies exploring the benefit of HRV modifications in CVD prevention are warranted.

## Background

Over the past decades, improved treatment and prevention of cardiovascular disease (CVD) have led to lower rates of ischemic events and better post-intervention outcomes in high-income countries [1]. However, the aging population is experiencing more vascular structural changes and cardiac remodeling [2] [3], which can lead to heart failure and subsequent lower quality of life and shorter life expectancy [4]. As acute cardiovascular events, heart failure, and early mortality are still major health concerns, we need to continue to improve early monitoring and identification of individuals with high risk of CVD.

People with a high risk of diabetes also have an increased risk of CVD and mortality [5, 6]. Subclinical indicators of diabetes-related microvascular and macrovascular complications can be present in individuals with pre-diabetes or high diabetes risk [7] [8]. As these people do not have a clinical diabetes diagnosis, they often remain outside of structured clinical management. Cardiovascular autonomic dysfunction (autonomic dysfunction), also known as cardiovascular autonomic neuropathy, can be detected in people with pre-diabetes and is pronounced with diabetes [9]. Autonomic dysfunction increases the risk of both CVD and mortality [10, 11]. Heart rate variability (HRV) is recognized as an indicator of cardiovascular autonomic function, as it quantifies the degree to which the sinoatrial node, which receives input from the autonomic nervous system can modulate the heart rate in response to various circumstances [12]. Studies have demonstrated that autonomic dysfunction (assessed by short period electrocardiograms (ECG)) is linked with CVD [10]. Fewer studies have investigated the association between long-term (> 24-hour) HRV and CVD [10]. Multiple days of HRV recording may capture an average of cardiovascular autonomic responses under regular free-living conditions that are less influenced by a person’s random activity during a particular day i.e. physical activity, emotion and sleep [13]. In addition, specific time-frames during circadian variation of HRV may be associated with CVD [14].

We aimed to determine the association between week-long HRV and the risk of incident CVD, heart failure, and all-cause mortality in a population with high risk of diabetes. Secondly, we wanted to identify the hours of the day with the strongest association between HRV and CVD, heart failure and all-cause mortality while accounting for the impact of concurrent physical acceleration and heart rate.

We hypothesized that 1) week-long HRV measures capture HRV patterns associated with risk of CVD events and that 2) the risk of CVD varies between hourly HRV measurements throughout the day.

## Methods

### Study population

Participants in the ADDITION-PRO prospective observational study were recruited between 2009 and 2011 from the Danish arm of the ADDITION-Europe study (ADDITION-DK) through a stepwise screening program for type 2 diabetes in primary care [15]. Ethical approval for the ADDITION-PRO study was obtained from the scientific ethics committee of the Central Denmark Region (Reference No. 20000183). The study was conducted in accordance with the Helsinki Declaration, and all participants provided oral and written informed consent for participation and for linkage of their data with national registers. ADDITION-PRO served as the follow-up health examination for individuals at high risk of developing diabetes [16]. The stratification of type 2 diabetes risk in ADDITION-DK was carried out using a Danish diabetes risk score questionnaire [15].

Participants were requested to report information about known risk factors for type 2 diabetes, including age, sex, BMI, known hypertension, family history of type 2 diabetes, gestational diabetes, and leisure time physical activity [17]. Those with a risk score of 5 points or more (out of a maximum of 15 points) were invited to attend a stepwise screening program, which included measures of random blood glucose levels and glycated hemoglobin A1c (HbA1c), a fasting blood glucose test (FPG), and an oral glucose tolerance test (OGTT). The World Health Organization criteria was utilized to diagnose type 2 diabetes [18]. The sampling frame for ADDITION-PRO included participants categorized into groups of increasing type 2 diabetes risk based on their diabetes risk score and glycemic status: low type 2 diabetes risk (less than 5 points on the diabetes risk score); high type 2 diabetes risk (5 or more points on the diabetes risk score) with normoglycemia, isolated impaired fasting glucose (IFG), isolated impaired glucose tolerance (IGT), or both IFG and IGT. A total of 2,082 individuals consented to participate in the ADDITION-PRO health examination, forming the baseline for this study [16]. Individuals with prior CVD events within 10 years before inclusion in ADDITION-PRO were excluded from this analysis. In the present study, we included participants with a valid HRV recording based on at least 48 hours of data and complete information on selected confounders, as described below.

### Heart Rate Variability

Heart rate was measured using a combination of an accelerometer and heart rate monitor (ActiHeart, CamNTech, Cambridge, UK). This monitor records uniaxial acceleration and heart rate [19]. The procedure of data collection and processing has been previously described [20]. From the ActiHeart, mean heart rates with prediction intervals were obtained every 30-second epoch. HRV is measured as the beat-to-beat variation between normal heart beat intervals on an electrocardiogram (ECG)[12]. Heart rate processing and calculations of HRV are fully described in supplemental material. Using the RHRV (version 4.2.7) package in R, we calculated HR and HRV indices [21]. We included the standard deviation between normal-to-normal heartbeat intervals (SDNN), the standard deviation of the 5-minute average NN intervals (SDANN), the SDNN index (SDNNi), and the triangular interpolation of NN interval histogram (TINN), and mean HR (mHR). The algorithm for these indices has been tested on a dataset with full 24-hour interbeat intervals (IBI), yielding high validity for global distributed HRV indices [22]. All HRV indices were calculated by week, 24-hour cycle, and for each hour of the day. As our data covered multiple days, we based our 24-hour and hour of the day indices on means across all cycles. HRV is dependent on individual baseline heart rate, e.g., an individual with higher resting heart rate will have less variation in heartbeat interval than an individual with lower heart rate.

Therefore, we pre-adjusted HRV for the heart rate at rest during the clinical visit. Resting pulse rate recordings at study visit were regressed on the logarithm (to obtain normality of residuals) to HRV. We then added the remaining residuals from the model to the intercept and transformed back into the original unit. As we wanted to test the impact of concurrent heart rate and physical acceleration on hourly HRV, we used the same method to pre-adjusted HRV for concurrent mHR and then further included physical acceleration in each particular hour.

### Outcomes

Information on CVD events and mortality, as well as all-cause mortality, was obtained from the Danish National Patient Registers until 2021. ICD-10 diagnosis codes for stroke, myocardial infarction, cardiovascular death, cardiovascular revascularization, and heart failure are described in supplementary material. We defined three outcomes: 1) three-point major adverse cardiovascular events (MACE), including fatal and non-fatal myocardial infarction, fatal and non-fatal stroke, cardiovascular revascularization, and cardiovascular death; 2) hospital-diagnosed heart failure; and 3) all-cause mortality.

### Covariates

Lifestyle factors, including smoking (current/ ex-smoker/ never) and alcohol consumption (average units per week), as well as CVD history and use of anti-hypertensive, glucose-lowering, and lipid-lowering medications, were obtained through a self-reported questionnaire. Physical activity energy expenditure kilojoule (KJ) per day (PAEE) was estimated based on combined accelerometry and heart rate data from ActiHeart recordings [20] and by the Recent Physical Activity Questionnaire (RPAQ). The hourly physical acceleration was based on accelerometer (m/s^2^) data alone. Blood measurements of HbA1c, OGTT, FPG, triglycerides, total cholesterol, high-density lipoprotein (HDL) cholesterol, and low-density lipoprotein (LDL) cholesterol were derived from blood samples. Body mass index (BMI), waist circumference, and systolic and diastolic blood pressure were measured during the participant’s clinical examination [16]. From the Danish registers, we collected information on CVD events in the 10 years prior to baseline and socioeconomic status at baseline (length of education, income, work status).

### Statistical Analysis

Baseline characteristics were described using mean and standard deviation (SD) for continuous variables and numbers (%) for categorical variables. Individual risk time was determined from the time-point of baseline data collection in ADDITION-PRO (2009-2011) until the time-point of CVD, death, or end of follow-up (31 December 2021).

### Analysis of week-long HRV

We used Poisson regression models to investigate the association between week-long SDNN and MACE, as well as hospitalized heart failure and all-cause mortality. SDNN and mHR were standardized by their mean and standard deviation. We fitted three models based on assumptions about potential confounding pathways summarized in directed acyclic graphs (DAG) (**Figure S1**). Model 1 included simple adjustments for age and sex. Model 2 accounted for confounding pathways visualized in a DAG (**Figure S1**), including further adjustment for education, alcohol consumption, smoking behavior, physical activity (PAEE calculated from RPAQ), body mass index, total cholesterol, and HbA1c. We used RPAQ to adjust for habitual physical activity and to avoid overadjustment from PAEE estimated from overlapping heart rate data. In model 3, we included systolic blood pressure, anti-hypertensive and glucose-lowering medications to account for use of medication and risk by elevated blood pressure. We performed analyses with HRV pre-adjusted for resting heart rate and included analyses of week-long mHR for comparison. To investigate non-linearity of the associations, we used splines, defining knots based percentiles in the HRV and mHR distribution. We performed additional analyses including HRV indices SDANN, SDNNi, TINN for the week-long recording and the mean across multiple 24-hour cycles of each HRV. These results are shown in the supplementary material (Table 2S and 3S).

### Analysis of Hourly HRV

We used Poisson regression models to investigate the association between hourly SDNN and MACE, hospitalization for heart failure, and all-cause mortality. We fitted two models based on assumptions from DAG (**Figure S2**). Each SDNN and mHR per hour was standardized by its mean and standard deviation. Model 1 included adjustments for age and sex. Model 2 was further adjusted for education, alcohol consumption, smoking behavior, PAEE (RPAQ calculated), body mass index, total cholesterol, and HbA1c. To test the influence of concurrent heart rate and physical acceleration, we performed analyses with mHR, physical acceleration, and heart rate and physical acceleration pre-adjusted HRV. All analyses were performed using multiple imputation chained equations to impute missing covariates, in the R statistical computing environment (version 4.2.2).

## Results

From the entire cohort, 1,627 (78%) participants had no prior CVD and week-long HRV measured and 1,432 had all hours represented with HRV with concurrent physical acceleration within a full-day (**Figure 1**). The study population included 53% men with a mean (SD) age of 66 (7) years and a mean BMI of 28 (5) kg/m^2^.

**Figure 1:**
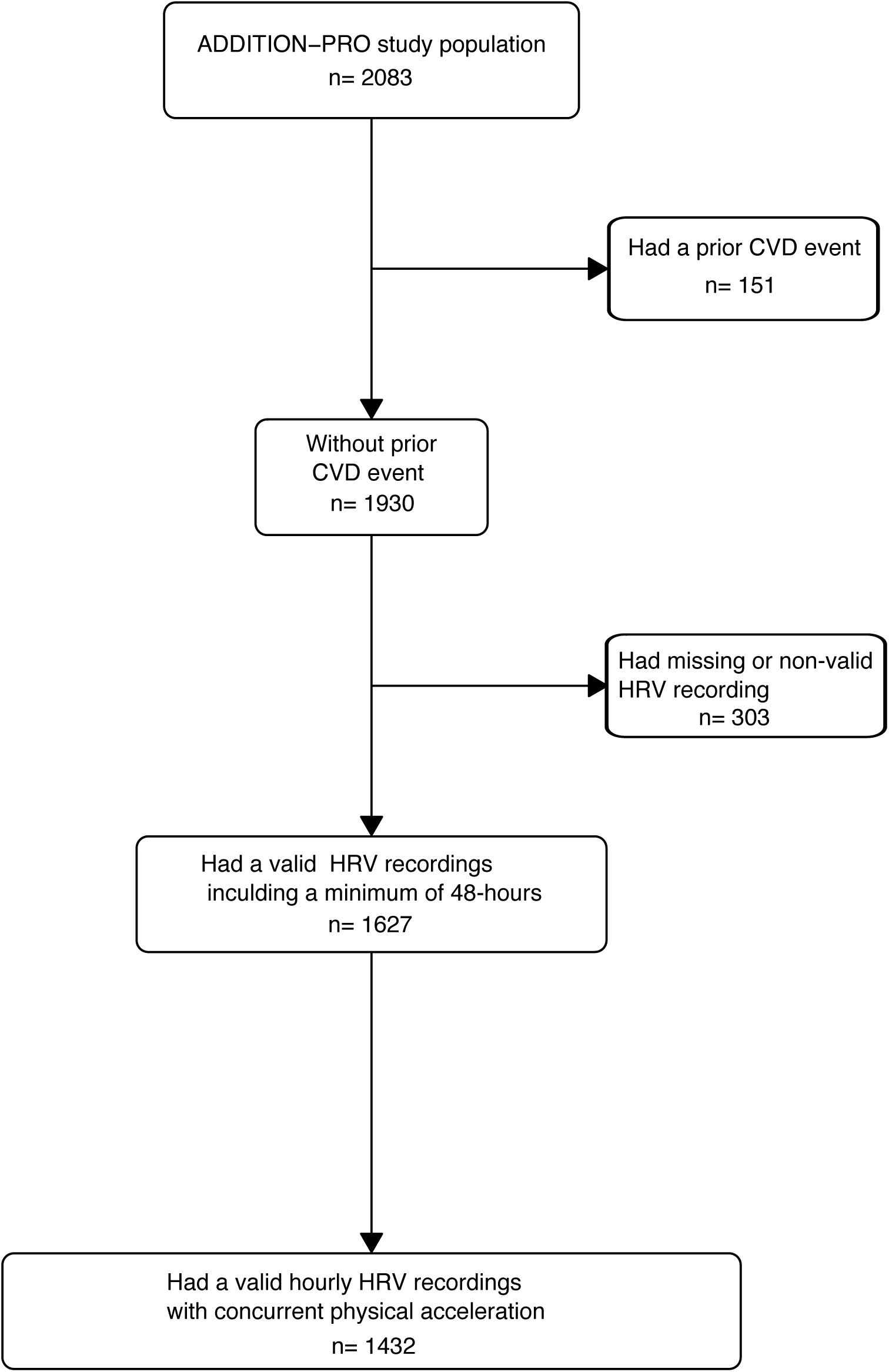
Study flowchart.

The mean week-long SDNN was 139.0 (32.3) ms and the heart rate was 73.5 (9.1) bpm. Hourly mean (SD) for SDNN, heart rate, and physical acceleration are presented in supplementary material (**Figure S2**). Forty-six percent had hypertension. Further characteristics of the participants are provided in **Table 1**. In total, the study population was followed for 17,926 person-years (Individual mean follow-up: 11.0 years). There were 173 incident cases of CVD (10.1 per 1000 person-years), 160 died (8.9 per 1000 person-years), and 71 received a hospital diagnosis of heart failure (4.0 per 1000 person-years).

**Table 1:** Baseline characteristics.

|  | Without CVD or HF event<br>N = 1,398 | With CVD or HF event<br>N = 227 |
| --- | --- | --- |
| Sex |  |  |
| Men | 722 (52%) | 144 (63%) |
| Women | 676 (48%) | 83 (37%) |
| Age (years) | 65.6 (6.9) | 68.0 (6.3) |
| Education (years) |  |  |
| <=10 years | 257 (19%) | 48 (21%) |
| >= 15 years | 450 (33%) | 63 (28%) |
| 10-15 years | 672 (49%) | 113 (50%) |
| Smoking status |  |  |
| Current | 225 (16%) | 38 (17%) |
| Never | 639 (46%) | 111 (49%) |
| Prior | 521 (38%) | 77 (34%) |
| Physical activity energy expenditure (KJ / day) | 53.2 (24.8) | 52.4 (26.5) |
| Alcohol consumption (units per week) | 9.1 (9.4) | 9.7 (9.6) |
| BMI (kg/m<sup>2</sup>) | 27.6 (4.7) | 28.2 (4.7) |
| Waist circumference (cm) | 96.4 (13.3) | 98.8 (13.6) |
| Systolic blood pressure (mmHg) | 133.2 (17.2) | 136.3 (17.7) |
| Diastolic blood pressure (mmHg) | 81.8 (10.3) | 82.6 (10.7) |
| Pulse rate (bpm) | 67.2 (10.7) | 68.2 (12.1) |
| HbA1c (%) | 5.8 (0.5) | 5.9 (0.6) |
| Triglycerides (mmol/L) | 1.3 (0.7) | 1.4 (0.8) |
| Total cholesterol (mmol/L) | 5.4 (1.1) | 5.3 (1.1) |
| HDL cholesterol (mmol/L) | 1.6 (0.4) | 1.5 (0.4) |
| LDL cholesterol (mmol/L) | 3.2 (1.0) | 3.2 (1.0) |
| Urine albumin-creatinine ratio (mg/g) | 22.6 (117.6) | 45.7 (201.6) |
| Use glucose lowering medication (yes) | 197 (14%) | 31 (14%) |
| Use anti-hypertensive medication (yes) | 617 (44%) | 136 (60%) |
| Use diuretics medication (yes) | 233 (17%) | 54 (24%) |
| Use beta blockers medication (yes) | 146 (11%) | 42 (19%) |
| Use calcium channel blockers medication (yes) | 183 (13%) | 53 (23%) |
| Use ACE inhibitors medication (yes) | 373 (27%) | 85 (38%) |
| Week-long standard deviation of all NN intervals (ms) | 139.6 (32.0) | 135.2 (33.6) |
| Week-long standard deviation of the averages of NN intervals in 5-minute segments (ms) | 116.5 (34.8) | 113.1 (36.0) |
| Week-long mean of the standard deviation for all 5 minutes segments (ms) | 54.8 (29.6) | 52.8 (27.7) |
| Week-long mean HR (bpm) | 73.4 (9.0) | 73.8 (9.8) |

n (%); Mean (SD)

### Week-long HRV index SDNN and mean heart rate association with major adverse cardiovascular events, heart failure and all-cause mortality

In our main analysis (model 2), 1 SD higher week-long SDNN was associated with a 0.82 (CI: 0.69; 0.97) incidence rate ratio (IRR) for MACE, 0.76 (CI: 0.58; 0.99) for heart failure and 0.79 (CI: 0.66; 0.94) for all-cause mortality (**Table 2**). Further adjustments for anti-hypertensive medication, systolic blood pressure, and pre-adjustment for resting heart rate did not materially change the estimates, except for heart failure where the association was partly attenuated. When examining association by splines, we observed nonlinear association with a higher IRR for MACE, heart failure, and all-cause mortality when SDNN was below approximately 120 ms (**Figure 2**). We observed no further reduction in IRR from levels of SDNN above 120 ms. The IRR of the whole week cycle was comparable with the mean of multiple 24-hour cycles of SDNN. Other HRV indices TINN, SDANN and SDNNi showed similar tendencies (**Table S2 and S3**).

**Figure 2:**
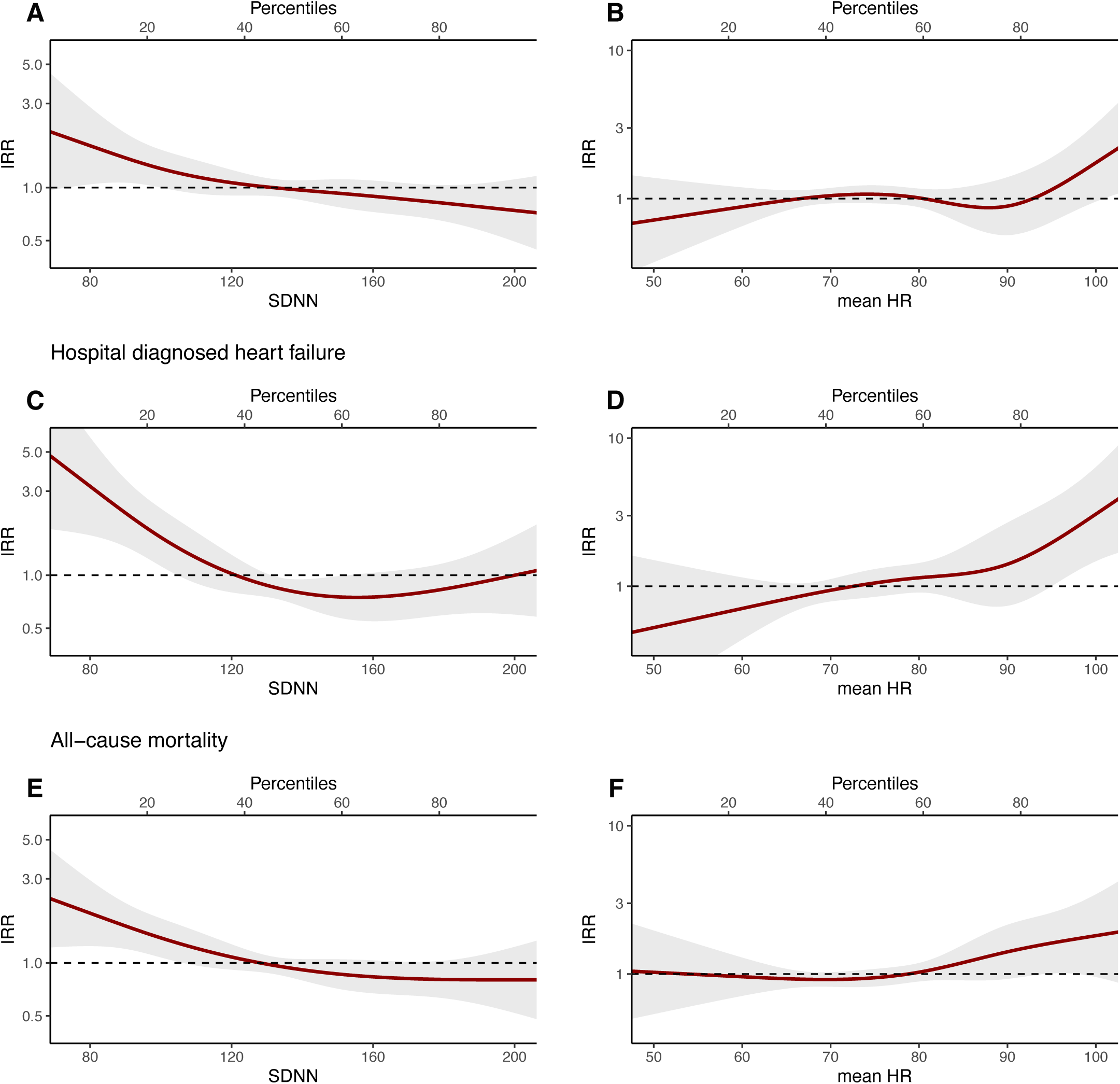
Week-long SDNN and mean HR association with major adverse cardiovascular events, heart failure, and all-cause mortality. Association between week-long HRV / mHR and MACE, hospital-diagnosed heart failure, and all-cause mortality. IRR are adjusted for age and sex, education, alcohol consumption, smoking behavior, physical activity, body mass index, total cholesterol, and Hba1c.

**Table 2:** Week-long SDNN and mean HR risk with Major adverse cardiovascular events, heart failure, and all-cause mortality.

|  | Model 1 | Model 2 | Model 3 |
| --- | --- | --- | --- |
|  | IRR (95% CI) | IRR (95% CI) | IRR (95% CI) |
| **Major adverse cardiovascular events** |  |  |  |
| SDNN | 0.79 (0.67; 0.93) | 0.82 (0.69; 0.97) | 0.83 (0.70; 0.98) |
| SDNN corrected for rHR | 0.82 (0.70; 0.96) | 0.85 (0.72; 1.00) | 0.86 (0.73; 1.01) |
| Mean HR | 1.11 (0.95; 1.29) | 1.07 (0.92; 1.25) | 1.08 (0.93; 1.26) |
| **Hospital diagnosed heart failure** |  |  |  |
| SDNN | 0.72 (0.56; 0.93) | 0.76 (0.58; 0.99) | 0.77 (0.59; 1.00) |
| SDNN corrected for rHR | 0.79 (0.62; 1.01) | 0.81 (0.63; 1.04) | 0.83 (0.65; 1.05) |
| Mean HR | 1.41 (1.14; 1.74) | 1.34 (1.07; 1.68) | 1.38 (1.10; 1.72) |
| **All-cause mortality** |  |  |  |
| SDNN | 0.69 (0.58; 0.82) | 0.79 (0.66; 0.94) | 0.80 (0.67; 0.95) |
| SDNN corrected for rHR | 0.75 (0.63; 0.88) | 0.84 (0.71; 0.99) | 0.85 (0.72; 1.00) |
| Mean HR | 1.23 (1.06; 1.42) | 1.12 (0.96; 1.31) | 1.14 (0.97; 1.32) |

*Incidence rate ratio for all-cause mortality and cardiovascular disease endpoints per SD in SDNN, HR corrected SDNN, and mHR over a week. Model 1: adjusted for age and sex; Model 2: model 1 + education, alcohol consumption, smoking behavior, physical activity, body mass index, total cholesterol, and Hba1c; Model 3: model 2 + systolic blood pressure, anti-hypertensive, and glucose-lowering medication.*

In model 2, 1 SD mHR was associated with a 1.34 (CI: 1.07; 1.68) higher IRR of heart failure. Week-long mHR did not show an association with MACE (1.07 [CI: 0.92; 1.25]) or all-cause mortality (1.12 [CI: 0.96; 1.31]). We observed a nonlinear association i.e. the IRR for heart failure and all-cause mortality were observed at mHR levels above 80 bpm (**Figure 2**). The threshold for higher risk of MACE was higher (92 bpm).

### Hourly SDNN and mean HR association with major adverse cardiovascular events, heart failure and all-cause mortality

When SDNN and mHR were divided into hourly segments during a day, we observed differences in risk with MACE, heart failure, and all-cause mortality across the 24-hours (**Figure 3**).

**Figure 3:**
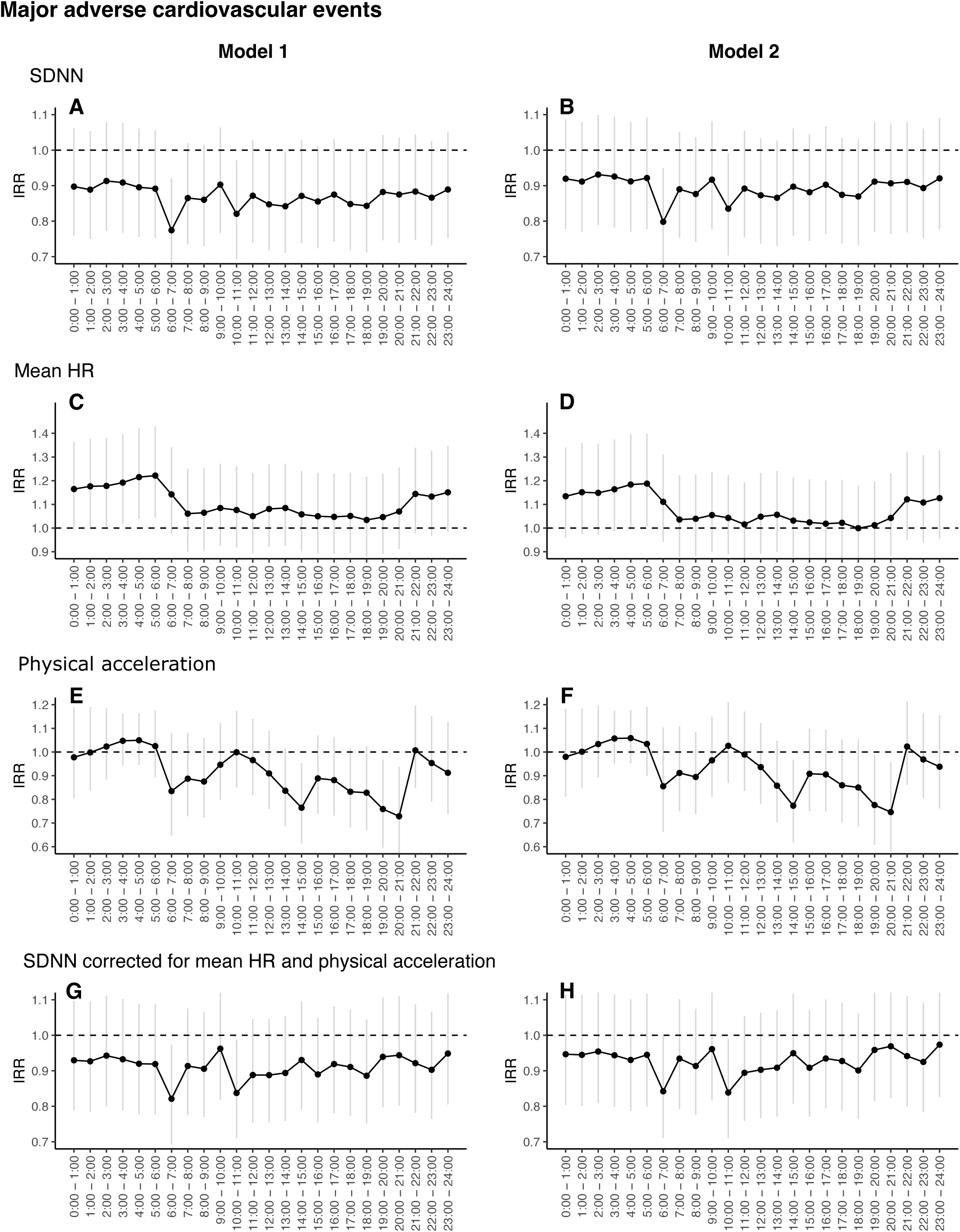
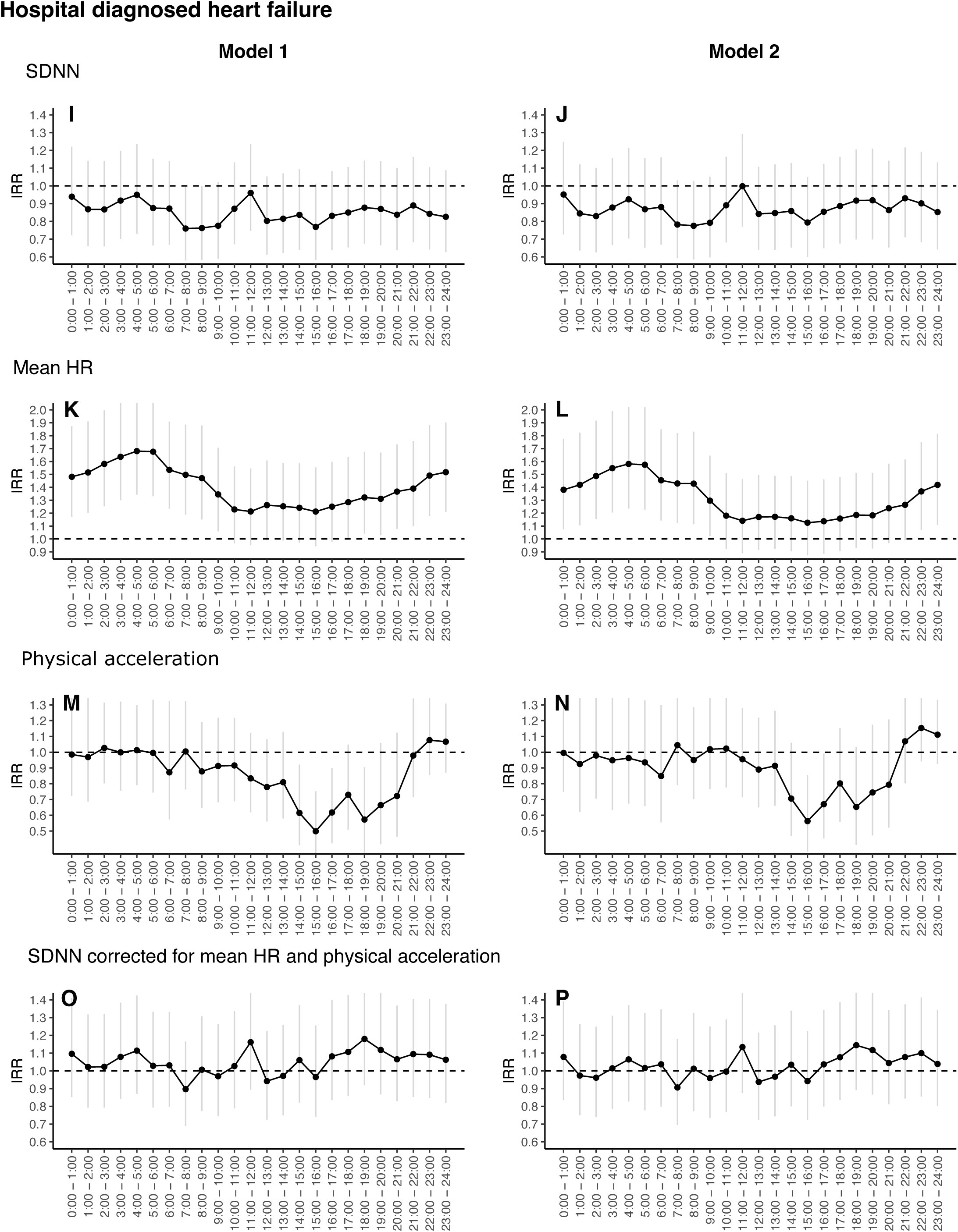
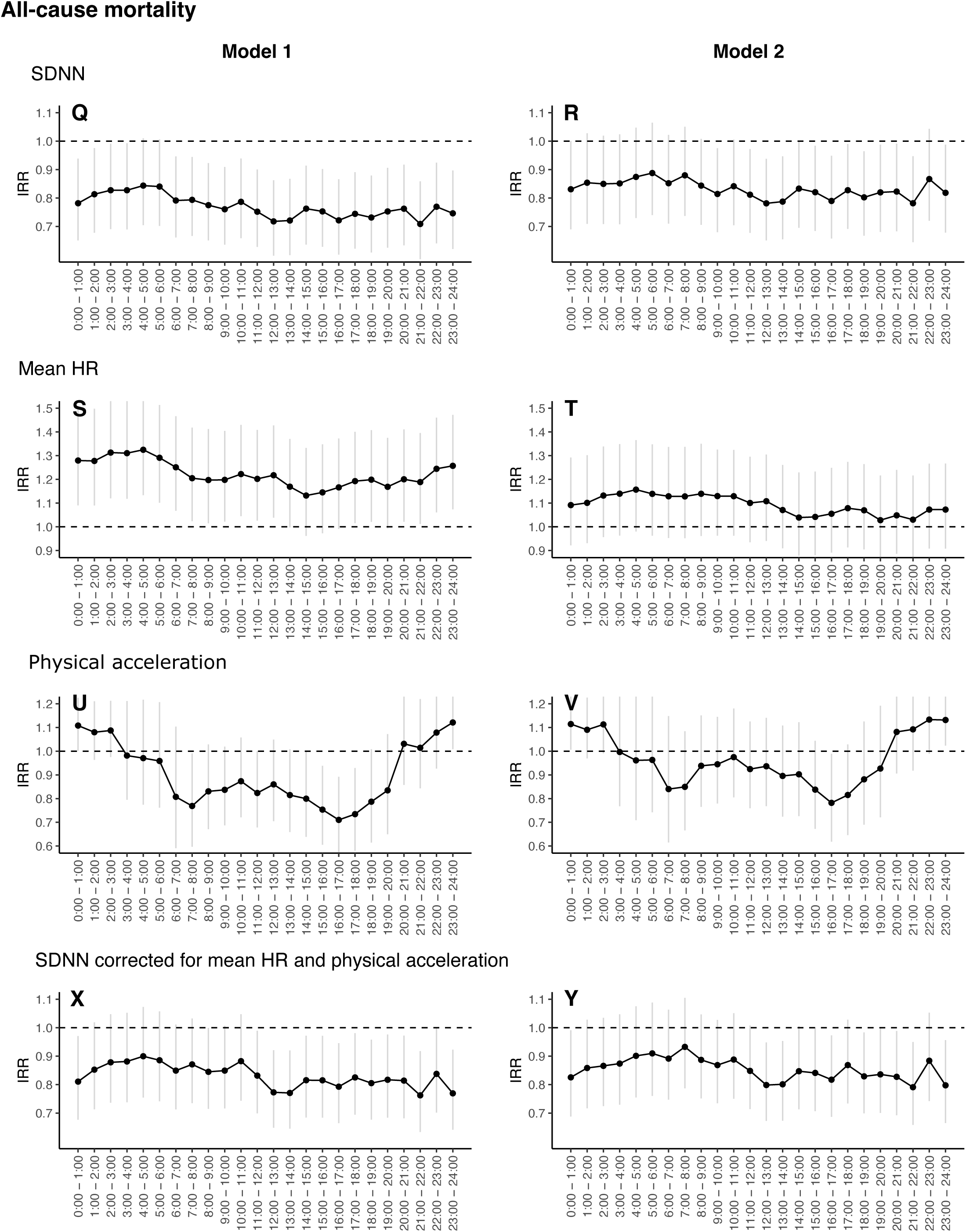
Diurnal heart rate variability and heart rate link with major adverse cardiovascular events, heart failure, and all-cause mortality risk. SDNN, mHR, physical acceleration, and preadjusted SDNN for concurrent physical acceleration and heart rate were measured each hour starting from 00:00 to 24:00. The figures (A-Y) are showing IRR of MACE, heart failure, and all-cause mortality per SD increase by each measurement across hour specific time-frames. Model 1: adjusted for age and sex; Model 2: model 1 + education, alcohol consumption, smoking behavior, body mass index, total cholesterol, and Hba1c.

#### Major adverse cardiovascular events

Per SD, across hourly time periods, SDNN showed similar lower risk of MACE, except for SDNN measured between 6:00 - 07:00 AM, which showed a lower adjusted IRR (0.80 [CI: 0.67; 0.95]) (**Figure 3B**). Pre-adjusting SDNN for concurrent mHR and physical acceleration slightly attenuated the results. mHR showed the strongest association with MACE between 04:00-06:00 AM e.g. per SD in bpm measured showed an adjusted IRR at 05:00-06:00 AM, 1.19 (CI: 1.01; 1.40).

#### Hospital-diagnosed heart failure

Measurements taken in the morning (from 7:00 - 10:00 AM) showed inverse associations between higher SDNN and IRR of heart failure. These trends were diminished after SDNN was preadjusted for concurrent physical acceleration and heart rate. Higher mHR measured during the night (from 2:00 - 5:00 AM) was associated with high IRR of heart failure (**Figure 3L**).

#### All-cause mortality

The association between higher SDNN and lower all-cause MRR was consistent across the 24 hours with a range of an adjusted MRR between 0.78 (CI: 0.65; 0.94) and 0.89 (CI: 0.74; 1.06). These associations were slightly attenuated after pre-adjustment for mHR and physical acceleration (**Figure 3Y**). mHR showed similar consistent trends association across 24-hours, where higher mHR was associated with higher risk of mortality (**Figure 3S**) but was attenuated after further adjustments.

## Discussion

Higher HRV index SDNN, assessed over a full week, is linked with an 18%, 24%, and 21% risk reduction per SD for MACE, heart failure, and all-cause mortality, respectively. The association showed a higher risk when SDNN values were below 120 ms. When the HRV periods were divided into hourly cycles, lower SDNN measured between 6:00-7:00 AM showed an association with a higher risk of MACE, whereas no particular time point had an exceptional association with all-cause MRR. Pre-adjusting hourly SDNN for concurrent physical acceleration and heart rate did not materially change the magnitude of these hourly associations. Also, higher mHR showed a higher risk of MACE and heart failure for measurements taken during the night hours from 02:00-06:00 AM.

We investigated week-long HRV in order to capture autonomic responses in free-living conditions over multiple days in a population with high risk of diabetes. Our findings showed a similar trend compared to 24-hour HRV in the association between HRV and CVD [23]. We consider it a strength to use recordings of HRV during multiple days and propose that such HRV measures are likely to contain more robust indications of individual autonomic responses to day-to-day situations. The trends of the association between the mean 24-hour HRV across multiple days compared to the complete week-long HRV were similar. Thus, both week-long and mean 24-hour HRV recordings can be used from the week-long measurement.

The link between autonomic dysfunction and both ischemic events and heart failure, may be attributed to an adverse cardiometabolic environment, and thus these risks are more pronounced in populations with high risk of diabetes and overt diabetes [24–26]. People with autonomic dysfunction, measured by a low week-long HRV, have a less adaptive autonomic nervous system response during the full day and night. Some of these dysadaptations seem to be more pronounced during specific hours of the day. Multiple explanations may contribute to explaining these associations.

Our results underscore the notion that autonomic dysfunction is not only linked with a high risk of CVD, but also a higher risk of all-cause mortality. Autonomic dysfunction might reflect a poor autonomic nervous adaptation, by parasympathetic impairment and sympathetic hyperresponsiveness, that affects adaptability in certain target organs e.g. the heart [27, 28]. Our results highlight that autonomic dysfunction in the morning, measured by lower HRV, is the strongest hourly indicator for higher risk of acute CVD endpoints and cardiovascular mortality. In the morning hours, the heart needs to make its biggest adaptation with the increase in sympathetic activity as the body experiences a peak in cortisol and changes from a longer rest to rise and movement [29]. Interestingly, lower HRV levels throughout the day (i.e. sleeping, waking, responding to physical movement, stress) are all indicative of mortality risk. The link between autonomic dysfunction with fatal and non-fatal CVD might be attributed to the arrhythmogenesis [28]. Why low HRV is linked with non-cardiovascular related death remains to be explored in future studies.

Lower week-long HRV might be an indicator of autonomic dysfunction driven by more chronic sympathetic dominance that leads to poorer deceleration of heart rate, changes of hemodynamics, and direct arterial constriction, and thus higher cardiac workload [30]. Hence, in the long term, autonomic dysfunction might cause pathological cardiac and arterial remodeling, which in turn increases the risk of ischemic events and heart failure [31, 32] [26]. Our findings show that higher heart rate in the night hours is a strong indicator for both heart failure, CVD, and mortality. These findings may highlight the absent parasympathetic activity during the sleeping/resting hours leading to minor changes in heart rate during rest. The explanation of the sympathetic overactivity is complex in whether the underlying cause is vagus nerve damage or a compensating sympathetic mechanism to keep sufficient ejection fraction [9, 33]. Therefore, we cannot exclude that higher heart rate might be an early indicator for progression to heart failure, as loss of stroke volume needs compensation by higher sympathetic activity, leading to higher heart rate that is needed throughout the day and night.

Recordings of long-term HRV are both influenced by habitual physical activity and actual physical activity during the measurement of HRV [13, 34]. Participants in ADDITION-PRO generally had low physical activity levels during the week-long recording [35]. When we included preadjusted hourly HRV for both heart rate and physical activity in the concurrent hour, the associations were slightly attenuated, but the trends of the associations were unchanged. Thus, we conclude that the association between higher HRV and lower CVD and mortality risk, is not solely explained by higher physical activity during the measurement time frame.

We prespecifed our DAG to close confounding pathways and avoid over-adjustments. The measurable confounding in the associations was mostly attributable to lifestyle factors, BMI, and biochemical markers. No material changes were observed when adding adjustments for systolic blood pressure, anti-hypertensive, and glucose-lowering medication. Pre-adjusting SDNN for baseline heart rate had a clearer impact on the IRR for heart failure, supported by the observed association between week-long mHR and heart failure. Thus, our findings suggest that heart rate serves as a marker of the development of heart failure, potentially reflecting either a predisposition due to a less healthy heart or the subtle progression of heart failure [36, 37]. The proportion explained between HRV and our outcomes showed that 25% of the SDNN association with heart failure was explained by baseline heart rate, compared to 14% for hard CVD events and 19% for mortality. Hence, this might underscore differences in the degree to which heart rate is an indicator of pre-clinical stages of the two outcomes. Therefore, we kept heart failure and the composite MACE as separate outcomes.

SDNN was included as our main determinant because it is the most frequently employed HRV index [12]. Participants in the ADDITION-PRO were invited based on their high risk of diabetes and have been followed up over a decade [16]. Therefore, our results highlight the potential use of measuring week-long heart rate and its variability for assessing incident CVD in populations with high risk of diabetes. Further studies are needed to determine whether these associations are valid in the general population, and in which risk and age groups HRV proves as a potential marker of CVD risk.

Our results demonstrate that in a population with a high risk of diabetes, assessing cardiovascular autonomic function may capture valuable knowledge of clinical relevance. In the current study population, the impact of one SD (33 ms) lower week-long SDNN was of equivalent magnitude to 4.5 additional years of aging for MACE risk and to 2.2-2.4 years for heart failure and all-cause mortality. Focus on cardiometabolic risk factor management could potentially lead to lower CVD and all-cause mortality risk by improving HRV. Findings support the notion but remain inconclusive due to the lack of trial evidence from drug or exercise interventions demonstrating a mediating role of HRV in the cardiometabolic prevention of CVD [38–41]. People living with high risk of diabetes can effectively modify their cardiometabolic risk profile through increased physical activity, which in addition can lead to improved autonomic function [42–44]. Therefore, HRV and heart rate are dynamic and responsive modifiable markers which potentially could be used to monitor potential successfulness in CVD risk management [14].

Over the past years, dynamic cardiovascular measures of heart rate and HRV have become more accessible by wearable devices [45]. Therefore, revisiting the use of these dynamic measures as a potential tool in risk management has become more relevant. Wearable devices have potential to improve health monitoring and facilitate targeted and individualized intervention based on physiological data [46, 47]. We showed that both long-term and hourly mHR and HRV contain relevant information about CVD and mortality risk. For example, morning HRV may represent an indicator for morning autonomic response that is stronger linked to MACE compared to the rest of the day response, and thus gives a notion for risk assessing time point in free living condition. Hence, monitoring heart rate and HRV could help us take early action when cardiovascular autonomic function deteriorates.

## Conclusion

Cardiovascular autonomic dysfunction, expressed by lower week-long HRV, is associated with higher risk of CVD and all-cause mortality in people with high risk of diabetes. There is heterogeneity in the associations across the hours of the day under free-living conditions that are not explained by physical acceleration and heart rate. Thus, long-term HRV and the diurnal autonomic response may capture different risks. Whether long-term HRV has potential as an effective modifiable marker or a prognostic indicator in higher risk populations, and where in the cardiovascular prevention trajectory it may play a role need further definition.

## Data Availability

The data that support the findings of this study are available from the Danish
Data Protection Agency and Danish Health Data Agency (j‑No: 2012‑58‑0004
and FSEID‑00002001) but restrictions apply to the availability of these data,
which were used under license for the current study, and so are not publicly
available. Data are however available from the authors upon reasonable
request and with permission of the Danish Data Protection Agency and Dan‑
ish Health Data Agency (j‑No: 2012‑58‑0004 and FSEID‑00002001).

## Abbreviations

CVD: Cardiovascular disease
MACE: Three-point major adverse cardiovascular events
ECG: Electrocardiogram
HRV: Heart rate variability
rHR: Resting heart rate
SDNN: The standard deviation of normal-to-normal R-R intervals
mHR: Mean heart rate
IRR: Incidence rate ratio
PAEE: Physical activity energy expenditure

## Acknowledgements

We would like to acknowledge all participants in the ADDITION-PRO, as well as research scientists, data managers and clinical and administrative staff who made the study possible. We would like to thank Luke W. Johnston for his expertise in data cleaning and processing, as well as Else-Marie Dalsgaard and Kasper Norman for their help with using Danish registries. We will also express our gratitude to Marianne Pedersen for her contribution as data manager on ADDITION-PRO.

## Authors’ contributions

Study concept and design: JRS, DRW, LB, DV, ELG, CSH. Contributed to the data: DRW, DV, AS. Planning the statistical analysis: JRS, DRW, LB. Conducted the statistical analysis: JRS, LB, DRW. All authors contributed to, critically revised, and approved the final version of the manuscript. JRS is the guarantor of this work and, as such, has full access to all the data in the study and takes responsibility for the integrity of the data and the accuracy of the data analysis.

## References

1. Timmis A, Kazakiewicz D, Torbica A, Townsend N, Huculeci R, Aboyans V, et al. Cardiovascular disease care and outcomes in West and South European countries. The Lancet Regional Health - Europe. 2023;33:100718. doi: 10.1016/j.lanepe.2023.100718.

2. Climie RE, Alastruey J, Mayer CC, Schwarz A, Laucyte-Cibulskiene A, Voicehovska J, et al. Vascular ageing: moving from bench towards bedside. European Journal of Preventive Cardiology. 2023;30(11):1101–17. doi: 10.1093/eurjpc/zwad028.

3. Gjesdal O, Bluemke DA, Lima JA. Cardiac remodeling at the population level—risk factors, screening, and outcomes. Nature Reviews Cardiology. 2011;8(12):673–85. doi: 10.1038/nrcardio.2011.154.

4. Conrad N, Judge A, Tran J, Mohseni H, Hedgecott D, Crespillo AP, et al. Temporal trends and patterns in heart failure incidence: a population-based study of 4 million individuals. The Lancet. 2018;391(10120):572–80. doi: 10.1016/S0140-6736(17)32520-5.

5. Birkenfeld AL, Franks PW, Mohan V. Precision Medicine in People at Risk for Diabetes and Atherosclerotic Cardiovascular Disease: A Fresh Perspective on Prevention. Circulation. 2024;150(24):1910–2. doi: doi:10.1161/CIRCULATIONAHA.124.070463.

6. Barr ELM, Zimmet PZ, Welborn TA, Jolley D, Magliano DJ, Dunstan DW, et al. Risk of Cardiovascular and All-Cause Mortality in Individuals With Diabetes Mellitus, Impaired Fasting Glucose, and Impaired Glucose Tolerance. Circulation. 2007;116(2):151–7. doi: doi:10.1161/CIRCULATIONAHA.106.685628.

7. Houben AJHM, Stehouwer CDA. Microvascular dysfunction: Determinants and treatment, with a focus on hyperglycemia. Endocrine and Metabolic Science. 2021;2:100073. doi: 10.1016/j.endmts.2020.100073.

8. Sörensen BM, Houben AJHM, Berendschot TTJM, Schouten JSAG, Kroon AA, van der Kallen CJH, et al. Prediabetes and Type 2 Diabetes Are Associated With Generalized Microvascular Dysfunction. Circulation. 2016;134(18):1339–52. doi: 10.1161/CIRCULATIONAHA.116.023446.

9. Coopmans C, Zhou TL, Henry RMA, Heijman J, Schaper NC, Koster A, et al. Both Prediabetes and Type 2 Diabetes Are Associated With Lower Heart Rate Variability: The Maastricht Study. Diabetes Care. 2020;43(5):1126–33. doi: 10.2337/dc19-2367.

10. Hillebrand S, Gast KB, de Mutsert R, Swenne CA, Jukema JW, Middeldorp S, et al. Heart rate variability and first cardiovascular event in populations without known cardiovascular disease: meta-analysis and dose–response meta-regression. EP Europace. 2013;15(5):742–9. doi: 10.1093/europace/eus341.

11. Jarczok MN, Weimer K, Braun C, Williams DP, Thayer JF, Gündel HO, et al. Heart rate variability in the prediction of mortality: A systematic review and meta-analysis of healthy and patient populations. Neuroscience & Biobehavioral Reviews. 2022;143:104907. doi: 10.1016/j.neubiorev.2022.104907.

12. Electrophysiology TFotESoCtNASoP. Heart Rate Variability. Circulation. 1996;93(5):1043–65. doi: doi:10.1161/01.CIR.93.5.1043.

13. Rietz M, Schmidt-Persson J, Gillies Banke Rasmussen M, Overgaard Sørensen S, Rath Mortensen S, Brage S, et al. Facilitating ambulatory heart rate variability analysis using accelerometry-based classifications of body position and self-reported sleep. Physiological Measurement. 2024.

14. Natarajan A, Pantelopoulos A, Emir-Farinas H, Natarajan P. Heart rate variability with photoplethysmography in 8 million individuals: a cross-sectional study. The Lancet Digital Health. 2020;2(12):e650–e7. doi: 10.1016/S2589-7500(20)30246-6.

15. Dalsgaard E-M, Christensen JO, Skriver MV, Borch-Johnsen K, Lauritzen T, Sandbaek A. Comparison of different stepwise screening strategies for type 2 diabetes: Finding from Danish general practice, Addition-DK. Primary Care Diabetes. 2010;4(4):223–9.

16. Johansen NB, Hansen AL, Jensen TM, Philipsen A, Rasmussen SS, Jørgensen ME, et al. Protocol for ADDITION-PRO: a longitudinal cohort study of the cardiovascular experience of individuals at high risk for diabetes recruited from Danish primary care. BMC Public Health. 2012;12:1078. doi: 10.1186/1471-2458-12-1078.

17. Christensen JO, Sandbaek A, Lauritzen T, Borch-Johnsen K. Population-based stepwise screening for unrecognised Type 2 diabetes is ineffective in general practice despite reliable algorithms. Diabetologia. 2004;47(9):1566–73. doi: 10.1007/s00125-004-1496-2.

18. Organization” WH. Definition, diagnosis and classification of diabetes mellitus and its complications: report of a WHO consultation. Part 1, Diagnosis and classification of diabetes mellitus. World health organization; 1999.

19. Brage S, Brage N, Franks PW, Ekelund U, Wareham NJ. Reliability and validity of the combined heart rate and movement sensor Actiheart. European Journal of Clinical Nutrition. 2005;59(4):561–70. doi: 10.1038/sj.ejcn.1602118.

20. Amadid H, Johansen NB, Bjerregaard AL, Brage S, Færch K, Lauritzen T, et al. The role of physical activity in the development of first cardiovascular disease event: a tree-structured survival analysis of the Danish ADDITION-PRO cohort. Cardiovasc Diabetol. 2018;17(1):126. doi: 10.1186/s12933-018-0769-x.

21. Martínez CAG, Quintana AO, Vila XA, Touriño MJL, Rodríguez-Liñares L, Presedo JMR, et al. Heart rate variability analysis with the R package RHRV. 2017.

22. Schaarup J: Actiheart validation of time-domain heart rate variability. https://figshare.com/articles/online_resource/Actiheart_validation_of_time-domain_heart_rate_variability/26182361 (2024). Accessed.

23. Wulsin LR, Horn PS, Perry JL, Massaro JM, D’Agostino RB. Autonomic Imbalance as a Predictor of Metabolic Risks, Cardiovascular Disease, Diabetes, and Mortality. J Clin Endocrinol Metab. 2015;100(6):2443–8. doi: 10.1210/jc.2015-1748.

24. Rinaldi E, van der Heide FCT, Bonora E, Trombetta M, Zusi C, Kroon AA, et al. Lower heart rate variability, an index of worse autonomic function, is associated with worse beta cell response to a glycemic load in vivo—The Maastricht Study. Cardiovascular Diabetology. 2023;22(1):105. doi: 10.1186/s12933-023-01837-0.

25. Schlaich M, Straznicky N, Lambert E, Lambert G. Metabolic syndrome: a sympathetic disease? Lancet Diabetes Endocrinol. 2015;3(2):148–57. doi: 10.1016/s2213-8587(14)70033-6.

26. Schaarup J, Bjerg L, Hansen C, Andersen S, Greevenbroek M, Schram M, et al. Cardiovascular autonomic dysfunction is linked with arterial stiffness across glucose metabolism: The Maastricht Study. 2024.

27. Rana S, Prabhu SD, Young ME. Chronobiological influence over cardiovascular function: the good, the bad, and the ugly. Circulation research. 2020;126(2):258–79.

28. Herring N, Kalla M, Paterson DJ. The autonomic nervous system and cardiac arrhythmias: current concepts and emerging therapies. Nature Reviews Cardiology. 2019;16(12):707–26. doi: 10.1038/s41569-019-0221-2.

29. Boudreau P, Dumont G, Kin NM, Walker CD, Boivin DB. Correlation of heart rate variability and circadian markers in humans. Annu Int Conf IEEE Eng Med Biol Soc. 2011;2011:681–2. doi: 10.1109/iembs.2011.6090153.

30. Nardone M, Floras JS, Millar PJ. Sympathetic neural modulation of arterial stiffness in humans. Am J Physiol Heart Circ Physiol. 2020;319(6):H1338–h46. doi: 10.1152/ajpheart.00734.2020.

31. Whelton SP, Blankstein R, Al-Mallah MH, Lima JAC, Bluemke DA, Hundley WG, et al. Association of Resting Heart Rate With Carotid and Aortic Arterial Stiffness. Hypertension. 2013;62(3):477–84. doi: doi:10.1161/HYPERTENSIONAHA.113.01605.

32. Schaarup JR, Christensen MS, Hulman A, Hansen CS, Vistisen D, Tabák AG, et al. Autonomic dysfunction is associated with the development of arterial stiffness: the Whitehall II cohort. GeroScience. 2023;45(4):2443–55. doi: 10.1007/s11357-023-00762-0.

33. Floras JS, Ponikowski P. The sympathetic/parasympathetic imbalance in heart failure with reduced ejection fraction. Eur Heart J. 2015;36(30):1974–82b. doi: 10.1093/eurheartj/ehv087.

34. Soares-Miranda L, Sattelmair J, Chaves P, Duncan GE, Siscovick DS, Stein PK, et al. Physical Activity and Heart Rate Variability in Older Adults. Circulation. 2014;129(21):2100–10. doi: doi:10.1161/CIRCULATIONAHA.113.005361.

35. Hansen A-LS, Carstensen B, Helge JW, Johansen NB, Gram B, Christiansen JS, et al. Combined Heart Rate– and Accelerometer-Assessed Physical Activity Energy Expenditure and Associations With Glucose Homeostasis Markers in a Population at High Risk of Developing Diabetes: The ADDITION-PRO study. Diabetes Care. 2013;36(10):3062–9. doi: 10.2337/dc12-2671.

36. Nanchen D, Leening MJG, Locatelli I, Cornuz J, Kors JA, Heeringa J, et al. Resting Heart Rate and the Risk of Heart Failure in Healthy Adults. Circulation: Heart Failure. 2013;6(3):403–10. doi: doi:10.1161/CIRCHEARTFAILURE.112.000171.

37. Ferrari R, Fox K. Heart rate reduction in coronary artery disease and heart failure. Nature Reviews Cardiology. 2016;13(8):493–501. doi: 10.1038/nrcardio.2016.84.

38. Tang Y, Shah H, Bueno Junior CR, Sun X, Mitri J, Sambataro M, et al. Intensive Risk Factor Management and Cardiovascular Autonomic Neuropathy in Type 2 Diabetes: The ACCORD Trial. Diabetes Care. 2020;44(1):164–73. doi: 10.2337/dc20-1842.

39. O’Driscoll JM, Wright SM, Taylor KA, Coleman DA, Sharma R, Wiles JD. Cardiac autonomic and left ventricular mechanics following high intensity interval training: a randomized crossover controlled study. Journal of Applied Physiology. 2018;125(4):1030–40. doi: 10.1152/japplphysiol.00056.2018.

40. Bönhof GJ, Strom A, Apostolopoulou M, Karusheva Y, Sarabhai T, Pesta D, et al. High-intensity interval training for 12 weeks improves cardiovascular autonomic function but not somatosensory nerve function and structure in overweight men with type 2 diabetes. Diabetologia. 2022;65(6):1048–57. doi: 10.1007/s00125-022-05674-w.

41. Andersen ST, Witte DR, Fleischer J, Andersen H, Lauritzen T, Jørgensen ME, et al. Risk Factors for the Presence and Progression of Cardiovascular Autonomic Neuropathy in Type 2 Diabetes: ADDITION-Denmark. Diabetes Care. 2018;41(12):2586–94. doi: 10.2337/dc18-1411.

42. Carnethon MR, Prineas RJ, Temprosa M, Zhang Z-M, Uwaifo G, Molitch ME, et al. The Association Among Autonomic Nervous System Function, Incident Diabetes, and Intervention Arm in the Diabetes Prevention Program. Diabetes Care. 2006;29(4):914–9. doi: 10.2337/diacare.29.04.06.dc05-1729.

43. Reduction in the Incidence of Type 2 Diabetes with Lifestyle Intervention or Metformin. New England Journal of Medicine. 2002;346(6):393–403. doi: doi:10.1056/NEJMoa012512.

44. Navarro-Lomas G, Dote-Montero M, Alcantara JMA, Plaza-Florido A, Castillo MJ, Amaro-Gahete FJ. Different exercise training modalities similarly improve heart rate variability in sedentary middle-aged adults: the FIT-AGEING randomized controlled trial. Eur J Appl Physiol. 2022;122(8):1863–74. doi: 10.1007/s00421-022-04957-9.

45. Dhingra LS, Aminorroaya A, Oikonomou EK, Nargesi AA, Wilson FP, Krumholz HM, et al. Use of Wearable Devices in Individuals With or at Risk for Cardiovascular Disease in the US, 2019 to 2020. JAMA Network Open. 2023;6(6):e2316634–e. doi: 10.1001/jamanetworkopen.2023.16634.

46. Schüssler-Fiorenza Rose SM, Contrepois K, Moneghetti KJ, Zhou W, Mishra T, Mataraso S, et al. A longitudinal big data approach for precision health. Nature Medicine. 2019;25(5):792–804. doi: 10.1038/s41591-019-0414-6.

47. Keshet A, Reicher L, Bar N, Segal E. Wearable and digital devices to monitor and treat metabolic diseases. Nature Metabolism. 2023;5(4):563–71. doi: 10.1038/s42255-023-00778-y.

