## supplemental material for "Cardiovascular autonomic dysfunction precedes cardiovascular disease and all-cause mortality: 11-year follow-up of the ADDITION-PRO study"

#### Heart rate variability based on 30-seconds mean heart rate and 95% prediction interval

We did not have access to the time series of successive normal to normal inter-beat intervals (IBI), also known as interbeat intervals (IBI), during the measurement period. Therefore, we generated random normal distribution IBIs for every 30-second interval based on the 30-second epoch of mean heart rate and prediction intervals. As earlier studies have shown that IBIs are normally distributed per 30-second epoch, we generated the IBI 30-second distribution using mean heart rate and standard deviation. To calculate SD from prediction intervals, we ensured that the prediction intervals differed symmetrically from the mean by calculating the difference between the upper and lower prediction intervals from the mean heart rate and visually observing their symmetry over time. Using the RHRV (version 4.2.7) package in R, we calculated HRV indices [1]. As we did not have successive time-series measurements, we only used HRV indices based on the distribution of RR intervals, available in time-domain and geometrical HRV indices [2].

**Reference:**

1. Martínez CAG, Quintana AO, Vila XA, Touriño MJL, Rodríguez-Liñares L, Presedo JMR, et al. Heart rate variability analysis with the R package RHRV. 2017.
2. Schaarup J: Actiheart validation of time-domain heart rate variability. https://figshare.com/articles/online_resource/Actiheart_validation_of_time-domain_heart_rate_variability/26182361 (2024). Accessed.

### Table S1: Diagnosis codes for of cardiovascular events

We defined CVD events by including ICD-10 diagnostic codes for stroke (ICD: DI61 - DI64) (SKA: KAAL10, KAAL11, KPAQ10, KPAQ20, KPAQ21), myocardial infarction (ICD: DI21-DI24), heart failure (ICD: DI50), and cardiovascular death (ICD: I20-I28, I42, I46, I50), and surgical codes for cardiovascular revascularization (SKA: KPAE10, KPAE25, KPAF10, KPAF20, KPAF21, KPAF22, KPAH10, KPAH20, KPAH21, KPEE, KPEF, KPEH, KPEP, KPEQ, KPFE, KPFH, KPFP, KPFQ).

| **Type of CVD event** | **Diagnosis codes** |
|---|---|
| Stroke | ICD: DI61 - DI64 |
| Myocardial infarction | ICD: DI21-DI24 |
| Heart failure | ICD: DI50 |
| Cardiovascular death | ICD: I20-I28, I42, I46, I50 |
| Cardiovascular revascularization | SKA: KPAE10, KPAE25, KPAF10, KPAF20, KPAF21, KPAF22, KPAH10, KPAH20, KPAH21, KPEE, KPEF, KPEH, KPEP, KPEQ, KPFE,, KPFH, KPFP, KPFQ |

### Table S2: Week-long HRV indices and mean HR association with major adverse cardiovascular events, heart failure, and all-cause mortality

| | Model 1 | Model 2 | Model 3 |
|---|---|---|---|
| | IRR (95% CI) | IRR (95% CI) | IRR (95% CI) |
| **Five-point MACE: AMI + Stroke + HF + All-cause mortality** | | | |
| SDNN | 0.80 (0.71; 0.89) | 0.85 (0.76; 0.96) | 0.86 (0.77; 0.97) |
| SDNN corrected for rHR | 0.84 (0.75; 0.94) | 0.89 (0.80; 1.00) | 0.90 (0.81; 1.01) |
| SDANN | 0.87 (0.78; 0.97) | 0.92 (0.82; 1.03) | 0.93 (0.83; 1.04) |
| SDANN corrected for rHR | 0.91 (0.82; 1.01) | 0.96 (0.86; 1.07) | 0.97 (0.87; 1.07) |
| SDNNIDX | 0.89 (0.80; 0.99) | 0.91 (0.82; 1.02) | 0.91 (0.81; 1.01) |
| SDNNIDX corrected for rHR | 0.92 (0.82; 1.02) | 0.93 (0.83; 1.03) | 0.92 (0.83; 1.03) |
| TINN | 0.81 (0.72; 0.90) | 0.87 (0.78; 0.97) | 0.87 (0.78; 0.98) |
| TINN corrected for rHR | 0.85 (0.76; 0.94) | 0.91 (0.81; 1.01) | 0.91 (0.82; 1.02) |
| Mean HR | 1.17 (1.05; 1.30) | 1.11 (1.00; 1.24) | 1.12 (1.01; 1.25) |
| **All-cause mortality** | | | |
| SDNN | 0.69 (0.58; 0.82) | 0.79 (0.66; 0.94) | 0.80 (0.67; 0.95) |
| SDNN corrected for rHR | 0.75 (0.63; 0.88) | 0.84 (0.71; 0.99) | 0.85 (0.72; 1.00) |
| SDANN | 0.80 (0.68; 0.94) | 0.91 (0.77; 1.07) | 0.92 (0.78; 1.08) |
| SDANN corrected for rHR | 0.85 (0.73; 1.00) | 0.94 (0.81; 1.10) | 0.95 (0.82; 1.12) |
| SDNNIDX | 0.82 (0.69; 0.97) | 0.85 (0.72; 1.00) | 0.84 (0.71; 1.00) |
| SDNNIDX corrected for rHR | 0.87 (0.73; 1.02) | 0.88 (0.75; 1.04) | 0.87 (0.74; 1.03) |
| TINN | 0.72 (0.61; 0.85) | 0.83 (0.70; 0.99) | 0.84 (0.71; 0.99) |
| TINN corrected for rHR | 0.77 (0.66; 0.90) | 0.88 (0.75; 1.03) | 0.88 (0.75; 1.04) |
| Mean HR | 1.23 (1.06; 1.42) | 1.12 (0.96; 1.31) | 1.14 (0.97; 1.32) |
| **Four-point MACE: AMI + Stroke + HF + CV Death** | | | |
| SDNN | 0.84 (0.73; 0.96) | 0.87 (0.75; 1.00) | 0.87 (0.76; 1.01) |
| SDNN corrected for rHR | 0.87 (0.76; 1.00) | 0.90 (0.78; 1.03) | 0.91 (0.79; 1.04) |
| SDANN | 0.89 (0.77; 1.01) | 0.91 (0.79; 1.05) | 0.92 (0.80; 1.06) |
| SDANN corrected for rHR | 0.92 (0.81; 1.06) | 0.95 (0.83; 1.09) | 0.95 (0.84; 1.09) |
| SDNNIDX | 0.92 (0.80; 1.05) | 0.93 (0.81; 1.06) | 0.93 (0.81; 1.06) |
| SDNNIDX corrected for rHR | 0.93 (0.82; 1.06) | 0.94 (0.82; 1.07) | 0.93 (0.82; 1.07) |
| TINN | 0.85 (0.74; 0.97) | 0.88 (0.76; 1.01) | 0.88 (0.77; 1.02) |
| TINN corrected for rHR | 0.89 (0.78; 1.01) | 0.91 (0.80; 1.05) | 0.92 (0.80; 1.05) |
| Mean HR | 1.13 (0.99; 1.28) | 1.10 (0.96; 1.26) | 1.11 (0.97; 1.27) |
| **Three-point MACE: AMI + Stroke + CV Death** | | | |
| SDNN | 0.79 (0.67; 0.93) | 0.82 (0.69; 0.97) | 0.83 (0.70; 0.98) |
| SDNN corrected for rHR | 0.82 (0.70; 0.96) | 0.85 (0.72; 1.00) | 0.86 (0.73; 1.01) |
| SDANN | 0.87 (0.75; 1.02) | 0.90 (0.77; 1.06) | 0.90 (0.77; 1.06) |
| SDANN corrected for rHR | 0.91 (0.78; 1.05) | 0.93 (0.80; 1.09) | 0.93 (0.80; 1.09) |
| SDNNIDX | 0.86 (0.73; 1.01) | 0.88 (0.75; 1.04) | 0.88 (0.75; 1.03) |
| SDNNIDX corrected for rHR | 0.88 (0.75; 1.03) | 0.90 (0.76; 1.05) | 0.89 (0.76; 1.05) |
| TINN | 0.83 (0.71; 0.97) | 0.86 (0.73; 1.01) | 0.87 (0.74; 1.02) |
| TINN corrected for rHR | 0.86 (0.74; 1.00) | 0.90 (0.76; 1.05) | 0.90 (0.77; 1.06) |
| Mean HR | 1.11 (0.95; 1.29) | 1.07 (0.92; 1.25) | 1.08 (0.93; 1.26) |

| | Model 1 | Model 2 | Model 3 |
|---|---|---|---|
| | IRR (95% CI) | IRR (95% CI) | IRR (95% CI) |
| **Hospital-diagnosed heart failure** | | | |
| SDNN | 0.72 (0.56; 0.93) | 0.76 (0.58; 0.99) | 0.77 (0.59; 1.00) |
| SDNN corrected for rHR | 0.79 (0.62; 1.01) | 0.81 (0.63; 1.04) | 0.83 (0.65; 1.05) |
| SDANN | 0.75 (0.59; 0.96) | 0.81 (0.63; 1.04) | 0.83 (0.64; 1.06) |
| SDANN corrected for rHR | 0.83 (0.65; 1.05) | 0.87 (0.68; 1.10) | 0.88 (0.70; 1.12) |
| SDNNIDX | 0.94 (0.74; 1.19) | 0.93 (0.73; 1.18) | 0.92 (0.72; 1.17) |
| SDNNIDX corrected for rHR | 0.96 (0.76; 1.22) | 0.93 (0.73; 1.18) | 0.92 (0.72; 1.17) |
| TINN | 0.68 (0.53; 0.87) | 0.72 (0.55; 0.93) | 0.72 (0.56; 0.93) |
| TINN corrected for rHR | 0.73 (0.57; 0.93) | 0.75 (0.59; 0.96) | 0.76 (0.60; 0.97) |
| Mean HR | 1.41 (1.14; 1.74) | 1.34 (1.07; 1.68) | 1.38 (1.10; 1.72) |

*Incidence rate ratio for all-cause mortality and cardiovascular disease endpoints per SD in SDNN, HR corrected SDNN, and mHR over a week. Model 1: adjusted for age and sex; Model 2: model 1 + education, alcohol consumption, smoking behavior, physical activity, body mass index, total cholesterol, and Hba1c; Model 3: model 2 + systolic blood pressure, anti-hypertensive, and glucose-lowering medication.*

### Table S3: Mean 24-hour HRV indices and mean HR association with major adverse cardiovascular events, heart failure, and all-cause mortality

| | Model 1 | Model 2 | Model 3 |
|---|---|---|---|
| | IRR (95% CI) | IRR (95% CI) | IRR (95% CI) |
| **Five-point MACE: AMI + Stroke + HF + All-cause mortality** | | | |
| SDNN | 0.80 (0.71; 0.89) | 0.85 (0.75; 0.95) | 0.85 (0.76; 0.96) |
| SDNN corrected for rHR | 0.83 (0.74; 0.92) | 0.87 (0.78; 0.97) | 0.88 (0.79; 0.98) |
| SDANN | 0.88 (0.79; 0.98) | 0.92 (0.83; 1.03) | 0.93 (0.83; 1.04) |
| SDANN corrected for rHR | 0.91 (0.81; 1.01) | 0.94 (0.85; 1.05) | 0.95 (0.86; 1.06) |
| SDNNIDX | 0.89 (0.80; 1.00) | 0.91 (0.82; 1.02) | 0.91 (0.81; 1.01) |
| SDNNIDX corrected for rHR | 0.92 (0.82; 1.02) | 0.93 (0.83; 1.03) | 0.92 (0.83; 1.03) |
| TINN | 0.79 (0.71; 0.88) | 0.84 (0.75; 0.95) | 0.85 (0.76; 0.95) |
| TINN corrected for rHR | 0.82 (0.74; 0.92) | 0.87 (0.78; 0.97) | 0.88 (0.78; 0.98) |
| Mean HR | 1.16 (1.05; 1.29) | 1.10 (0.99; 1.23) | 1.11 (1.00; 1.24) |
| **All-cause mortality** | | | |
| SDNN | 0.71 (0.60; 0.84) | 0.80 (0.68; 0.96) | 0.81 (0.68; 0.96) |
| SDNN corrected for rHR | 0.75 (0.63; 0.88) | 0.83 (0.70; 0.98) | 0.84 (0.71; 0.99) |
| SDANN | 0.84 (0.71; 0.98) | 0.93 (0.79; 1.10) | 0.94 (0.80; 1.11) |
| SDANN corrected for rHR | 0.87 (0.74; 1.01) | 0.95 (0.81; 1.11) | 0.96 (0.82; 1.12) |
| SDNNIDX | 0.81 (0.68; 0.96) | 0.84 (0.71; 1.00) | 0.83 (0.70; 0.99) |
| SDNNIDX corrected for rHR | 0.85 (0.72; 1.01) | 0.87 (0.73; 1.03) | 0.86 (0.73; 1.02) |
| TINN | 0.72 (0.61; 0.85) | 0.82 (0.69; 0.97) | 0.82 (0.69; 0.98) |
| TINN corrected for rHR | 0.76 (0.65; 0.90) | 0.85 (0.72; 1.00) | 0.85 (0.72; 1.01) |
| Mean HR | 1.22 (1.06; 1.41) | 1.11 (0.95; 1.30) | 1.12 (0.96; 1.31) |
| **Four-point MACE: AMI + Stroke + HF + CV Death** | | | |
| SDNN | 0.82 (0.71; 0.94) | 0.85 (0.73; 0.98) | 0.85 (0.74; 0.98) |
| SDNN corrected for rHR | 0.85 (0.74; 0.98) | 0.87 (0.76; 1.00) | 0.88 (0.77; 1.01) |
| SDANN | 0.87 (0.76; 1.00) | 0.90 (0.78; 1.03) | 0.90 (0.79; 1.04) |
| SDANN corrected for rHR | 0.91 (0.79; 1.04) | 0.93 (0.81; 1.06) | 0.93 (0.82; 1.06) |
| SDNNIDX | 0.93 (0.81; 1.06) | 0.94 (0.82; 1.07) | 0.94 (0.82; 1.07) |
| SDNNIDX corrected for rHR | 0.94 (0.83; 1.08) | 0.95 (0.83; 1.08) | 0.94 (0.83; 1.08) |
| TINN | 0.82 (0.71; 0.94) | 0.84 (0.73; 0.97) | 0.85 (0.73; 0.98) |
| TINN corrected for rHR | 0.84 (0.74; 0.97) | 0.87 (0.75; 1.00) | 0.87 (0.76; 1.00) |
| Mean HR | 1.12 (0.98; 1.27) | 1.09 (0.95; 1.25) | 1.10 (0.96; 1.26) |
| **Three-point MACE: AMI + Stroke + CV Death** | | | |
| SDNN | 0.79 (0.67; 0.93) | 0.82 (0.69; 0.97) | 0.83 (0.70; 0.98) |
| SDNN corrected for rHR | 0.82 (0.70; 0.96) | 0.85 (0.72; 0.99) | 0.86 (0.73; 1.00) |
| SDANN | 0.88 (0.76; 1.03) | 0.91 (0.77; 1.06) | 0.91 (0.78; 1.07) |
| SDANN corrected for rHR | 0.91 (0.79; 1.06) | 0.93 (0.80; 1.09) | 0.94 (0.80; 1.09) |
| SDNNIDX | 0.87 (0.74; 1.02) | 0.89 (0.76; 1.04) | 0.89 (0.76; 1.04) |
| SDNNIDX corrected for rHR | 0.89 (0.76; 1.04) | 0.90 (0.77; 1.06) | 0.90 (0.77; 1.05) |
| TINN | 0.80 (0.69; 0.94) | 0.84 (0.71; 0.99) | 0.84 (0.71; 0.99) |
| TINN corrected for rHR | 0.83 (0.71; 0.97) | 0.86 (0.74; 1.01) | 0.87 (0.74; 1.02) |
| Mean HR | 1.11 (0.95; 1.29) | 1.07 (0.92; 1.25) | 1.08 (0.92; 1.26) |

| | Model 1 | Model 2 | Model 3 |
|---|---|---|---|
| | IRR (95% CI) | IRR (95% CI) | IRR (95% CI) |
| **Hospital diagnosed heart failure** | | | |
| SDNN | 0.67 (0.52; 0.86) | 0.70 (0.54; 0.92) | 0.71 (0.55; 0.92) |
| SDNN corrected for rHR | 0.72 (0.57; 0.92) | 0.74 (0.57; 0.95) | 0.75 (0.59; 0.96) |
| SDANN | 0.70 (0.55; 0.90) | 0.76 (0.58; 0.98) | 0.77 (0.60; 0.99) |
| SDANN corrected for rHR | 0.76 (0.60; 0.97) | 0.80 (0.63; 1.02) | 0.81 (0.64; 1.03) |
| SDNNIDX | 0.95 (0.76; 1.20) | 0.95 (0.75; 1.20) | 0.94 (0.74; 1.19) |
| SDNNIDX corrected for rHR | 0.98 (0.78; 1.23) | 0.95 (0.75; 1.20) | 0.94 (0.74; 1.19) |
| TINN | 0.64 (0.49; 0.82) | 0.67 (0.51; 0.87) | 0.67 (0.51; 0.87) |
| TINN corrected for rHR | 0.67 (0.52; 0.86) | 0.69 (0.54; 0.89) | 0.69 (0.54; 0.90) |
| Mean HR | 1.37 (1.10; 1.70) | 1.31 (1.04; 1.63) | 1.34 (1.07; 1.67) |

*Incidence rate ratio for all-cause mortality and cardiovascular disease endpoints per SD in SDNN, HR corrected SDNN, and mHR over a week. Model 1: adjusted for age and sex; Model 2: model 1 + education, alcohol consumption, smoking behavior, physical activity, body mass index, total cholesterol, and Hba1c; Model 3: model 2 + systolic blood pressure, anti-hypertensive, and glucose-lowering medication.*

**Figure S1: DAG1 – Week-long HRV and CVD, heart failure, and all-cause mortality**

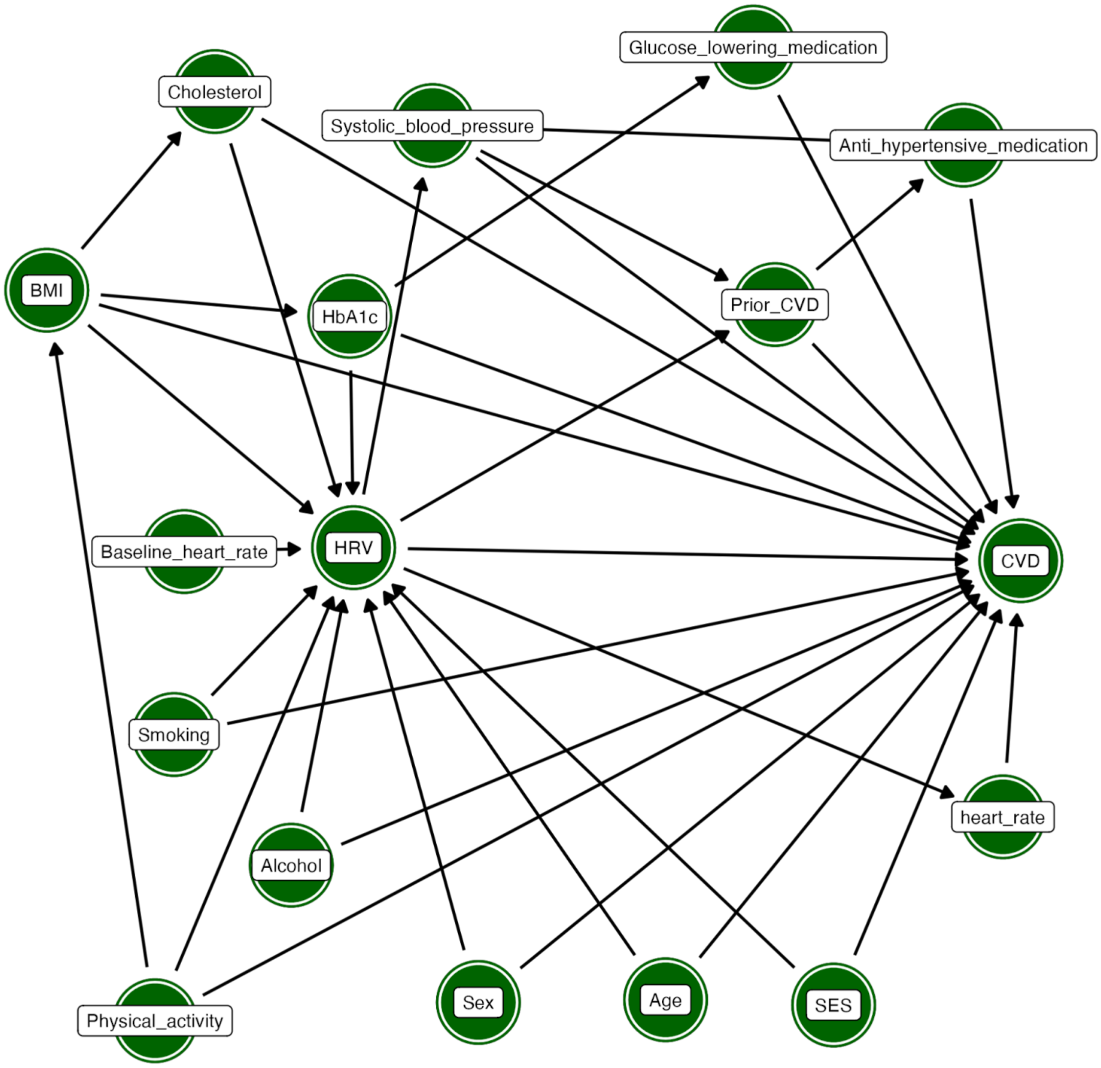

Confounding pathways *in the first study aim* vizualized by *directed acyclic graphs (DAG). Aim: To determine the risk between week-long HRV and CVD, heart failure, and all-cause mortality in a population with high-risk of diabetes.*

**Figure S2: DAG2 – Hourly HRV and CVD, heart failure, and all-cause mortality**

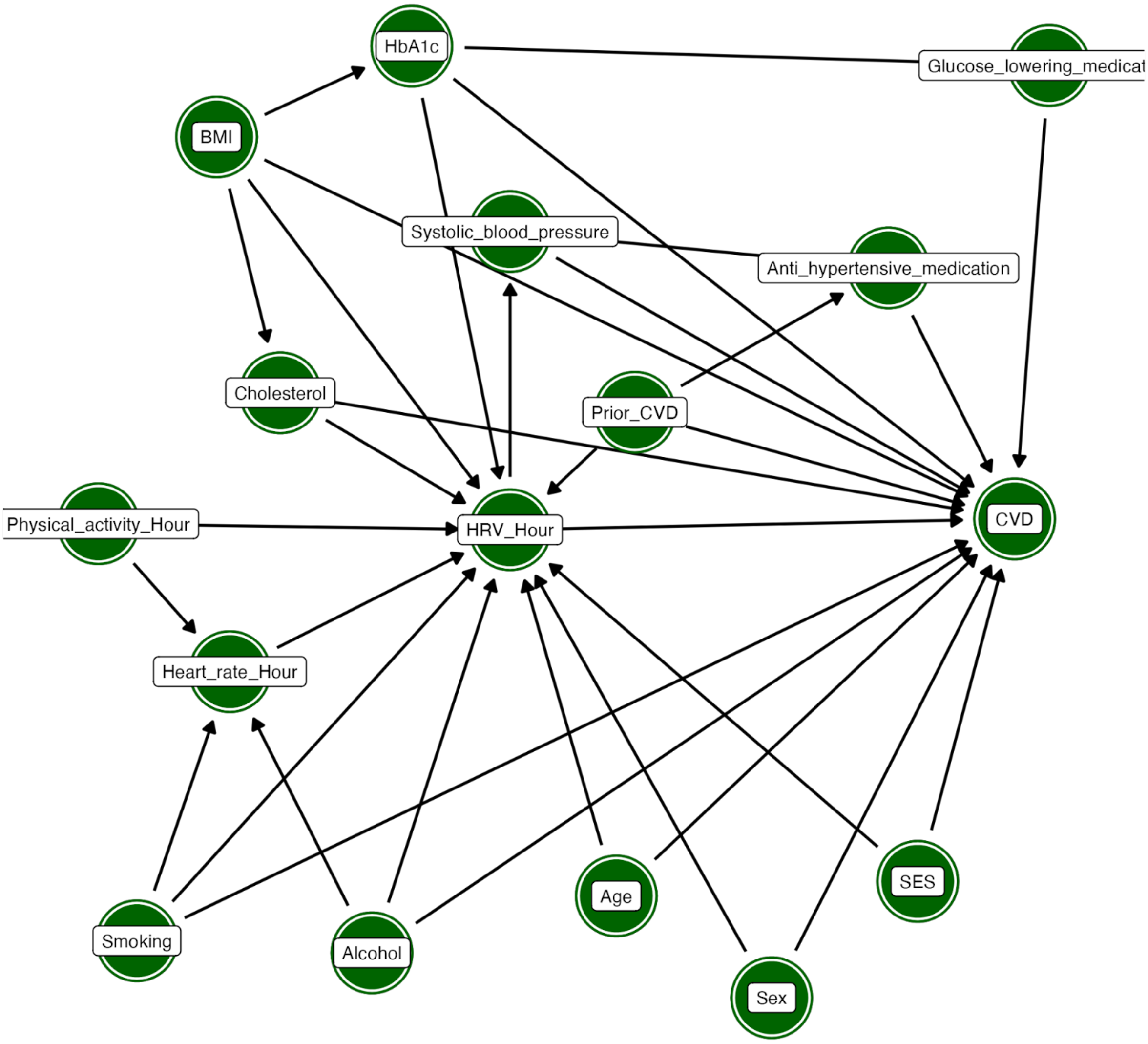

Confounding pathways *in the second study aim* vizualized by *directed acyclic graphs(DAG). Aim: To identify the hours of the day where HRV has the strongest association with CVD, heart failure, and all-cause mortality risk and test the impact of concurrent physical acceleration and heart rate.*

**Figure S3: Hourly SDNN, heart rate, physical acceleration and sleep over 24 hours**

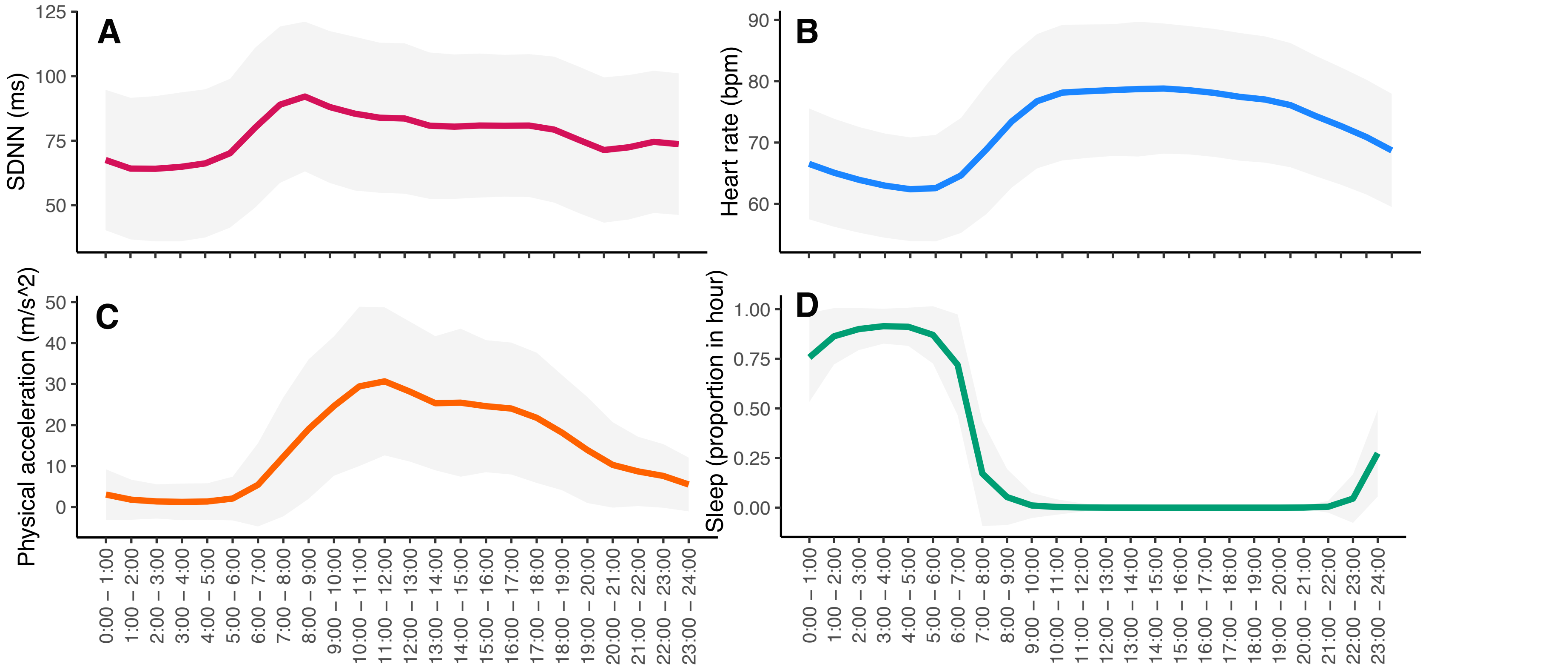

Mean and standard deviation of SDNN (A), heart rate (B), physical acceleration (C), and sleep (D) in each hour time-frame across 24-hours.

### Figure S4: Week-long SDNN corrected for rHR association with MACE, heart failure, and all-cause mortality

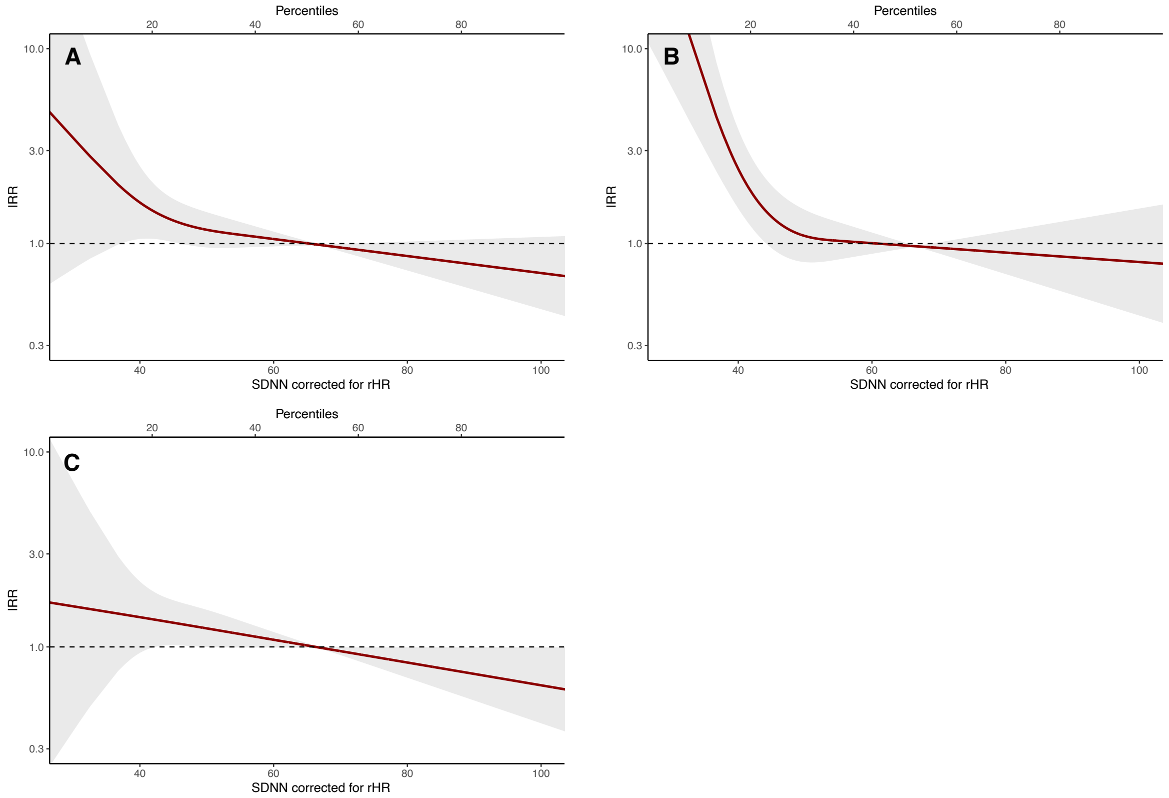

*Association between week-long SDNN corrected for rHR and MACE (A), hospital-diagnosed heart failure (B), and all-cause mortality (C). IRR are adjusted for age and sex, education, alcohol consumption, smoking behavior, physical activity, body mass index, total cholesterol, and Hba1c.*
